# Systematic review of Economic studies of Partner Notification and management interventions for sexually transmitted infections including HIV in men who have sex with men

**DOI:** 10.1101/2021.06.08.21258534

**Authors:** CB Okeke Ogwulu, Z Abdali, EV Williams, CS Estcourt, AR Howarth, A Copas, F Mapp, M Woode-Owusu, TE Roberts

## Abstract

**Objectives:** Men who have sex with men (MSM) are disproportionately affected by sexually transmitted infections (STIs). Partner notification (PN) to identify, test and treat sex partners of MSM with bacterial STIs is challenging because MSM often report larger numbers of sex partners and a higher proportion of one-off partners who may be difficult to engage. However, one-off partners contribute disproportionately to onward transmission. Economic research on PN has typically focused on heterosexual people and evidence of effectiveness of PN in MSM is scant. We conducted a systematic review of economic studies of PN interventions in MSM to inform the development of a novel PN intervention for MSM with one-off partners.

**Method:** Six electronic databases were searched up to June 2020. Cost studies and full economic evaluations, which focused on PN and/or testing and treatment (in the context of PN) of sex partners of MSM with STIs, and/or HIV, were included. A two-stage categorisation process was used for study selection and a narrative synthesis was reported.

**Results:** Twenty-six studies of a possible 1909 met the selection criteria. Sixteen focused on MSM but only three of these were on PN. Few studies reported on patients’ characteristics and settings. Most studies were cost-utility analyses with outcomes reported as quality-adjusted life years (QALYs) which were derived from studies on heterosexual people.

**Conclusions:** None of the identified studies specifically addressed cost-effectiveness of PN in MSM. The few studies identified as potentially relevant relied on costs and QALYs data from studies in heterosexual people, which may be inappropriate given the different patterns of sexual partnerships reported by these two groups. The lack of evidence on efficient PN approaches for MSM, a group with a high burden of infection, supports the need for new interventions tailored to the needs and preferences of MSM with parallel economic evaluation.

## INTRODUCTION

Men who have sex with men (MSM) bear a disproportionate burden of sexually transmitted infections (STIs) and HIV. Among MSM with STIs, the pattern of sexual partners tends to differ from heterosexuals and is characterised by higher numbers of partners and a greater proportion of one-off partners who contribute disproportionately to community transmission.^1^

Partner notification (PN), the process of identifying, testing and treating exposed sex partners is a key element of STI control.^2^ However, few studies have focussed on PN among MSM and achieving even modest outcomes is challenging.^3^ This is of concern because of (a) increasing antimicrobial resistance to Neisseria gonorrhoea (the causative agent of gonorrhoea), with MSM accounting for the majority of cases^4^ and (b) failure to reach MSM exposed to an STI wastes opportunities for health promotion and engagement with newer interventions to reduce the risk of other STIs and blood-borne viruses such as, vaccination for Human papillomavirus (HPV) and HIV Pre Exposure prophylaxis (PrEP). The differences in numbers and patterns of sexual partners reported by MSM mean that different PN interventions may be needed and that the resources required to achieve good PN outcomes may be greater than for heterosexuals.

As part of work to develop novel PN interventions for MSM.^5^ we conducted a systematic review of economic studies of PN interventions for STIs in MSM to ground future intervention development in economic reality. PN involves testing and often treatment of sex partners. Hence, to ensure comprehensive inclusion of all PN-related components of care, we also explored studies associated with testing and treatment of MSM with STIs as the settings and methods of engagement of the population for testing and treatment may provide insights on how to target PN for MSM.

## METHODS

A systematic review was conducted following the UK’s Centre for Review and Dissemination (CRD) guidelines and reported according to the Preferred Reporting Items for Systematic Reviews and Meta-Analyses (PRISMA) guidelines where appropriate.^6^ A comprehensive search strategy (*S1*) was formulated using the PICO framework.

Following a scoping search on Google Scholar and MEDLINE, six electronic databases including - MEDLINE, EMBASE, Web of Science, NHS Economic Evaluation Database (NHS EED), HIMC and CINAHL – were searched up to June 2020 by CO and ZA. The reference lists of potentially key papers were hand-searched to identify additional papers. Search results were entered into the endnote database manager, to exclude irrelevant studies and code relevant studies.

Inclusion and exclusion criteria (*S2*) were applied. A two-stage process (*S3*) was used to screen studies for inclusion using published methods.^7^ Categorisation was conducted independently by two reviewers (CO and ZA). No formal quality appraisal was applied given the aim of inclusivity of relevant information.

## RESULTS

Twenty-six studies were selected for inclusion in the narrative synthesis (*S4*). All studies were conducted in high-income countries and included 22 formal economic evaluations and four cost studies. The studies focused on MSM (16), heterosexuals (8), or heterosexuals and MSM (1). One study focused on males but did not indicate their sexual behaviour. Fifteen studies reported on either HIV only (11) or HIV and STIs (4). The remaining 11 studies were on Chlamydia (6) Gonorrhea (3) and one each on Syphilis and Hepatitis B Virus (HBV) infection.

Overall, 11 studies has a primary focus on PN (with or without testing/treatment). The remaining studies were on testing (10), treatment (3), both treatment and testing (2) (*Table 1 and S5)*. We present a primary narrative synthesis of the PN papers with additional information from other studies presented in *S6*.

**Table 1:**
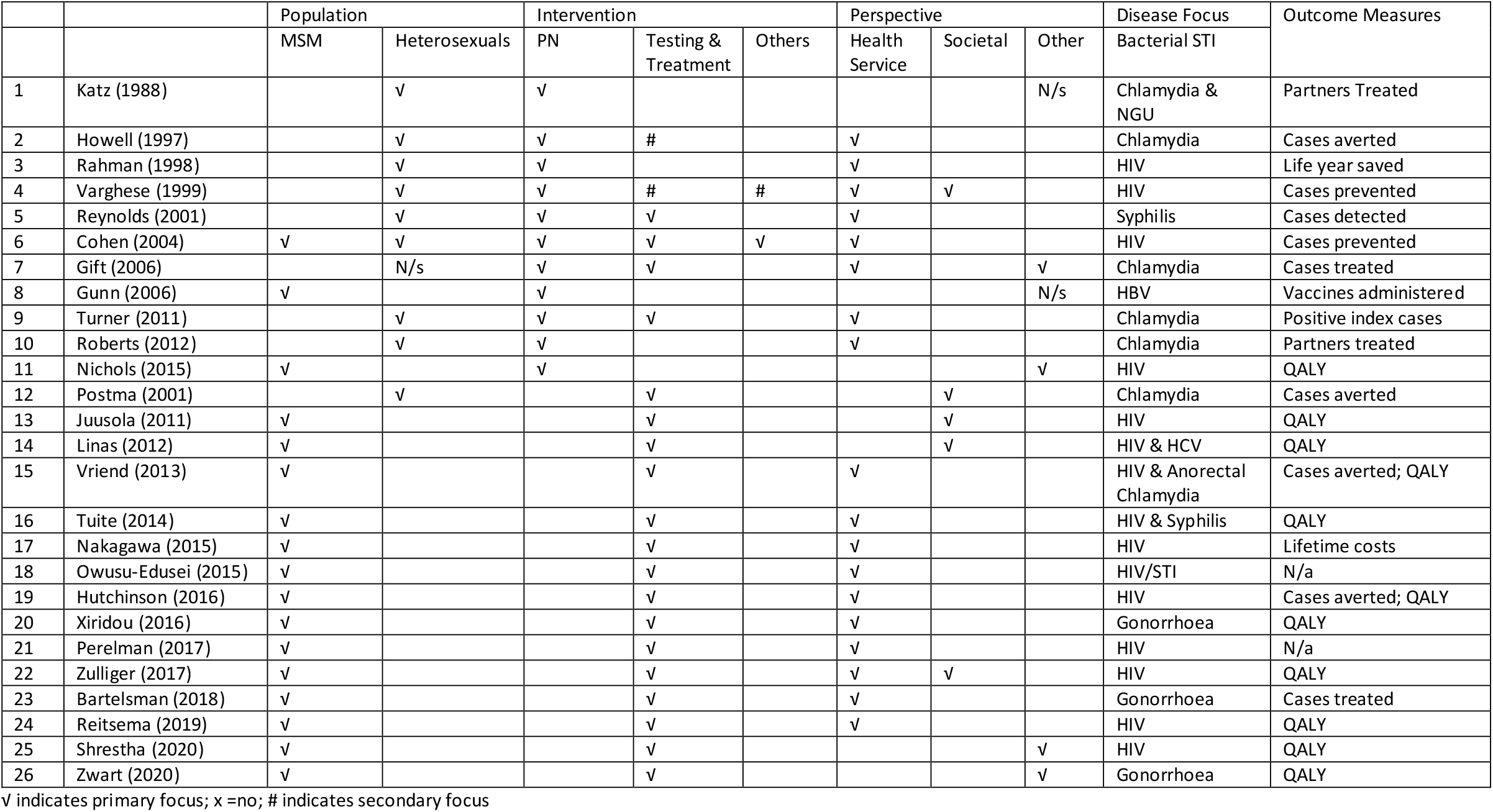
Summary of Results

|  |  | Population |  | Intervention |  |  | Perspective |  |  | Disease Focus | Outcome Measures |
| --- | --- | --- | --- | --- | --- | --- | --- | --- | --- | --- | --- |
|  |  | MSM | Heterosexuals | PN | Testing & Treatment | Others | Health Service | Societal | Other | Bacterial STI |  |
| 1 | Katz (1988) |  | √ | √ |  |  |  |  | N/s | Chlamydia & NGU | Partners Treated |
| 2 | Howell (1997) |  | √ | √ | # |  | √ |  |  | Chlamydia | Cases averted |
| 3 | Rahman (1998) |  | √ | √ |  |  | √ |  |  | HIV | Life year saved |
| 4 | Varghese (1999) |  | √ | √ | # | # | √ | √ |  | HIV | Cases prevented |
| 5 | Reynolds (2001) |  | √ | √ | √ |  | √ |  |  | Syphilis | Cases detected |
| 6 | Cohen (2004) | √ | √ | √ | √ | √ | √ |  |  | HIV | Cases prevented |
| 7 | Gift (2006) |  | N/s | √ | √ |  | √ |  | √ | Chlamydia | Cases treated |
| 8 | Gunn (2006) | √ |  | √ |  |  |  |  | N/s | HBV | Vaccines administered |
| 9 | Turner (2011) |  | √ | √ | √ |  | √ |  |  | Chlamydia | Positive index cases |
| 10 | Roberts (2012) |  | √ | √ |  |  | √ |  |  | Chlamydia | Partners treated |
| 11 | Nichols (2015) | √ |  | √ |  |  |  |  | √ | HIV | QALY |
| 12 | Postma (2001) |  | √ |  | √ |  |  | √ |  | Chlamydia | Cases averted |
| 13 | Juusola (2011) | √ |  |  | √ |  |  | √ |  | HIV | QALY |
| 14 | Linas (2012) | √ |  |  | √ |  |  | √ |  | HIV & HCV | QALY |
| 15 | Vriend (2013) | √ |  |  | √ |  | √ |  |  | HIV & Anorectal Chlamydia | Cases averted; QALY |
| 16 | Tuite (2014) | √ |  |  | √ |  | √ |  |  | HIV & Syphilis | QALY |
| 17 | Nakagawa (2015) | √ |  |  | √ |  | √ |  |  | HIV | Lifetime costs |
| 18 | Owusu-Edusei (2015) | √ |  |  | √ |  | √ |  |  | HIV/STI | N/a |
| 19 | Hutchinson (2016) | √ |  |  | √ |  | √ |  |  | HIV | Cases averted; QALY |
| 20 | Xiridou (2016) | √ |  |  | √ |  | √ |  |  | Gonorrhoea | QALY |
| 21 | Perelman (2017) | √ |  |  | √ |  | √ |  |  | HIV | N/a |
| 22 | Zulliger (2017) | √ |  |  | √ |  | √ | √ |  | HIV | QALY |
| 23 | Bartelsman (2018) | √ |  |  | √ |  | √ |  |  | Gonorrhoea | Cases treated |
| 24 | Reitsema (2019) | √ |  |  | √ |  | √ |  |  | HIV | QALY |
| 25 | Shrestha (2020) | √ |  |  | √ |  |  |  | √ | HIV | QALY |
| 26 | Zwart (2020) | √ |  |  | √ |  |  |  | √ | Gonorrhoea | QALY |

√ indicates primary focus; x=no; # indicates secondary focus

### Partner Notification Studies

Eleven studies assessed PN interventions of which three involved MSM. Two were solely among MSM and one included MSM and heterosexuals. The disease focus of the three studies was HIV and HBV. The remaining eight studies were among heterosexuals with disease focus on chlamydia, HIV and syphilis (*Table 1 and S7*).

### Patient Characteristics

Most PN studies used secondary data with limited reporting of patient characteristics. Of the three MSM papers, Gunn et al. ^8^evaluated the usefulness of a Syphilis PN model amongst 129 MSM and injecting drug users (IDU) with chronic HBV aged 15-45 years. The study concluded that PN was limited by the high proportion of MSM index cases with anonymous partners or expressing a preference to inform their own partners.

Nichols et al. ^9^ used a hypothetical (aged ≥ 15 years) cohort to represent the Dutch HIV epidemic among MSM. The authors used outcome data from a Public Health Service in The Netherlands where the PN strategy was supported by an online PN tool, Suggest-A-Test.

Cohen et al ^10^., assessed the relative cost-effectiveness of 26 HIV prevention strategies including PN to change risk behaviours of MSM and heterosexuals in the US. The authors assessed patterns of cost-effectiveness across different populations. For MSM, they concluded the most cost-effective interventions were individually focused interventions such as PN and counselling. None of these studies reported on the types of partners such as regular or one-off partners.

### Economic Evaluations and outcomes

The 11 PN papers included one CEA/CUA, eight CEAs, one CCA and one cost analysis (*S7 and S8*). Six of the CEA studies incorporated decision trees, or mathematical models. (used to predict long-term and final outcomes). Most papers were not explicit about the rationale for the models. The CEAs reported outcomes as natural units including cost per: patients treated; cases averted; cases detected; or partner treated which are typically intermediate outcomes for STIs.

Nichols et al. conducted a model-based CEA/CUA to predict the long-term effectiveness of PN for MSM with HIV and reported outcomes as cost per quality-adjusted life-years (QALYs) gained with QALY weights from a published meta-analysis of HIV/AIDS utility estimateswhich were derived from heterosexual men and women. They assessed two PN scenarios versus no PN with a time horizon of 5 years and 20 years. The authors concluded that PN was cost-effective in the short-term and that its cost-effectiveness increases over time.

Cohen et al., developed a spreadsheet tool that incorporated a mathematical model based on the Bernoulli process used by previous studies. The authors reported outcomes as cost per new HIV infection prevented including primary infections (directly prevented by the intervention) and secondary infections prevented in sex partners. Gunn et al. presented a cost analysis of PN services and reported results as PN services cost per vaccine.

### Perspectives and costs

Only one paper adopted the societal perspective, recognised as the gold standard because it includes all costs (direct and indirect healthcare; direct non-healthcare) and consequences borne by society. The paper focused on heterosexuals with HIV and collected resource use data for direct medical costs and costs incurred by clients and the provider. Six studies including two of the MSM studies adopted the perspective of the healthcare system or provider and reported direct medical costs. The remaining papers were not explicit on the perspective adopted but collected data on direct medical costs only.

The PN papers provided results on average or total costs for a range of indices with one exception. Cohen et al explored MSM and PN but included heterosexuals. However, total costs for PN were only provided for the heterosexual population in this study. Gunn et al. conducted a cost analysis of HBV vaccine for MSM (*S6*).

## DISCUSSION

We found very little evidence on the health economic aspects of PN interventions among MSM. Two studies focussed on PN for HIV, one on HBV and no studies considered PN for bacterial STIs. Amongst the PN studies, there was one CUA which reported outcomes in terms of cost per QALYs. However, the QALYs estimates were from previous studies on heterosexuals. Consequently, the true applicability of the results to the MSM population is questionable.

To our knowledge, this is the first systematic review focussing on health economic aspects of PN in MSM. Findings provide some useful costs information on PN for STIs/HIV in this group that could be used as model inputs for future analyses but the lack of studies on bacterial STIs is notable.

Given the paucity of economic studies and the burden of STIs and HIV in MSM, more research is needed to inform development of new PN interventions tailored to this group’s needs. This must include health economic considerations. In particular, the possibility of eliciting utility values (for QALYs) from the MSM population. One-off partners, who may contribute disproportionately to onward transmission might benefit from digital PN options^5^ which appear particularly under-studied. While we understand the need to resort to data from other population in the absence of evidence, this suggests there is currently no robust evidence to support any particular approach. The reviewed studies do not provide enough information on PN amongst MSM; hence, we cannot draw conclusions on outcomes and cost-effectiveness of PN for this population.

## Data Availability

Not applicable

