## Supplemental S1 for "Systematic review of Economic studies of Partner Notification and management interventions for sexually transmitted infections including HIV in men who have sex with men": Supplementary Information.pdf

### *S1: Keywords and MESH terms used for the search*

| |
|---|
| 1. HIV/ or HIV.mp. or HIV-2/ or HIV-1/ |
| 2. Human Immunodeficiency virus.mp. or HIV/ |
| 3. AIDS.mp. or Acquired Immunodeficiency Syndrome/ |
| 4. Acquired Immunodeficiency Syndrome/ or HIV Infections/ or HIV Aids.mp. |
| 5. STI.mp. or Sexually Transmitted Diseases/ |
| 6. STD.mp. or Sexually Transmitted Diseases/ |
| 7. Chlamydia Infections/ or Gonorrhea/ or Sexually Transmitted Diseases/ or sexually transmitted*.mp. or HIV Infections/ |
| 8. chlamydia*.mp. or Chlamydia Infections/ or Chlamydia/ or Chlamydia trachomatis/ |
| 9. Gonorrhea/ or gonorrhoea.mp. or Neisseria gonorrhoea/ |
| 10. Syphilis/ or syphillis.mp. |
| 11. Hepatitis C/ or Hepatitis/ or Hepatitis B/ |
| 12. Lymphogranuloma Venereum/ or LGV.mp. |
| 13. 1 or 2 or 3 or 4 or 5 or 6 or 7 or 8 or 9 or 10 or 11 or 12 |
| 14. Homosexuality, Male/ or MSM.mp. or Risk-Taking/ or Sexual Behavior/ |
| 15. men who have sex with men.mp. or "Sexual and Gender Minorities"/ |
| 16. Gay.mp. or "Sexual and Gender Minorities"/ |
| 17. bisexual.mp. or "Sexual and Gender Minorities"/ |
| 18. Homosexuality/ or homosexual*.mp. or Homosexuality, Male/ |
| 19. 14 or 15 or 16 or 17 or 18 |
| 20. test*.mp. |
| 21. treat*.mp. |
| 22. screen*.mp. |
| 23. partner notification.mp. or Contact Tracing/ |
| 24. PN.mp. |
| 25. contact tracing.mp. or Contact Tracing/ |
| 26. 20 or 21 or 22 or 23 or 24 or 25 |
| 27. 13 and 19 and 26 |
| 28. economic evaluation.mp. or Cost-Benefit Analysis/ |
| 29. costs.mp. or "Costs and Cost Analysis"/ |
| 30. "Costs and Cost Analysis"/ or cost studies.mp. or Cost-Benefit Analysis/ or "Cost of Illness"/ |
| 31. Health Care Costs/ or "Costs and Cost Analysis"/ or Cost-Benefit Analysis/ or cost* analysis.mp. |
| 32. cost effectiveness analysis.mp. or Cost-Benefit Analysis/ |
| 33. cost utility analysis.mp. or Cost-Benefit Analysis/ |
| 34. cost consequence analysis.mp. or Health Care Costs/ |
| 35. cost minimisation analysis.mp. or Hospital Costs/ |
| 36. Models, Economic/ or economic model*.mp. |
| 37. modeling.mp. or Patient-Specific Modeling/ |
| 38. 28 or 29 or 30 or 31 or 32 or 33 or 34 or 35 or 36 or 37 |
| 39. 27 and 38 |
| 40. limit 39 to (english language and humans) |

### *S2: Inclusion and Exclusion Criteria*

***Participants***: MSM with STIs including bacterial STIs: Chlamydia, Gonorrhoea and Syphilis and/or HIV.

***Intervention***: PN, testing or treatment strategies in the context of partner management.

***Outcomes***: Economic input and output data.

***Study*** type: Formal economic evaluations conducted in any setting, for any STI including HIV, including cost-effectiveness analysis (CEA), cost-utility analysis (CUA) and cost-consequences analysis (CCA); other cost studies including cost analysis published in English.

There were no restrictions on year or country. Papers that described PN interventions in other populations were explored for possible generalisability of evidence to the MSM population. Studies that reported on the use of technology for interventions not related to PN such as health promotion and education were excluded.

#### *S3: Selection of Studies*

A two-stage process was used to screen studies for inclusion using published methods.

**Stage 1: Initial Categorisation of studies**

Each study was categorised independently by two reviewers (CO and ZA) into Categories A-F based on title and abstract where available.

The study reports primary or secondary research on the costs or utilisation of PN in MSM (STI and HIV), and includes formal economic evaluation.

The study reports primary or secondary research on the costs or utilisation of testing and treatment in MSM (STI and HIV), and includes formal economic evaluation.

The study reports primary or secondary research on the costs or utilisation of PN **i**n STIs and HIV and includes formal economic evaluation.

The study reports primary or secondary research on the costs or utilisation of, testing and treatment in STIs and HIV, and includes formal economic evaluation.

The study may have useful information on PN but does not specifically fall into A to D.

The study does not have any relevance to the economic evaluation of interventions for PN.

Papers in Categories A to C were considered relevant to the systematic review. A random sample of 25% of the articles in category (D) were retrieved and reviewed in full. If 20% of these articles are considered useful then the remaining 75% of the articles in category (D) will be retrieved and reviewed in full, otherwise, all papers in category (D) will be excluded from the review.

**Stage 2: Further categorisation of studies**

For Stage 2, full texts of studies in groups A, B and C were read to classify them further using the following criteria:

Formal Economic evaluation (a) UK or high-income studies (b) LMICs

Other cost studies a) UK or high-income studies (b) LMICs

Effectiveness study, with some assessment of implications for cost or quantity of resources used.

Not relevant to the economic evaluation of interventions for PN in MSM

All articles classified as (A1), (A2), (B1), (B2), (C1) or (C2) were included in the next stage of the review.

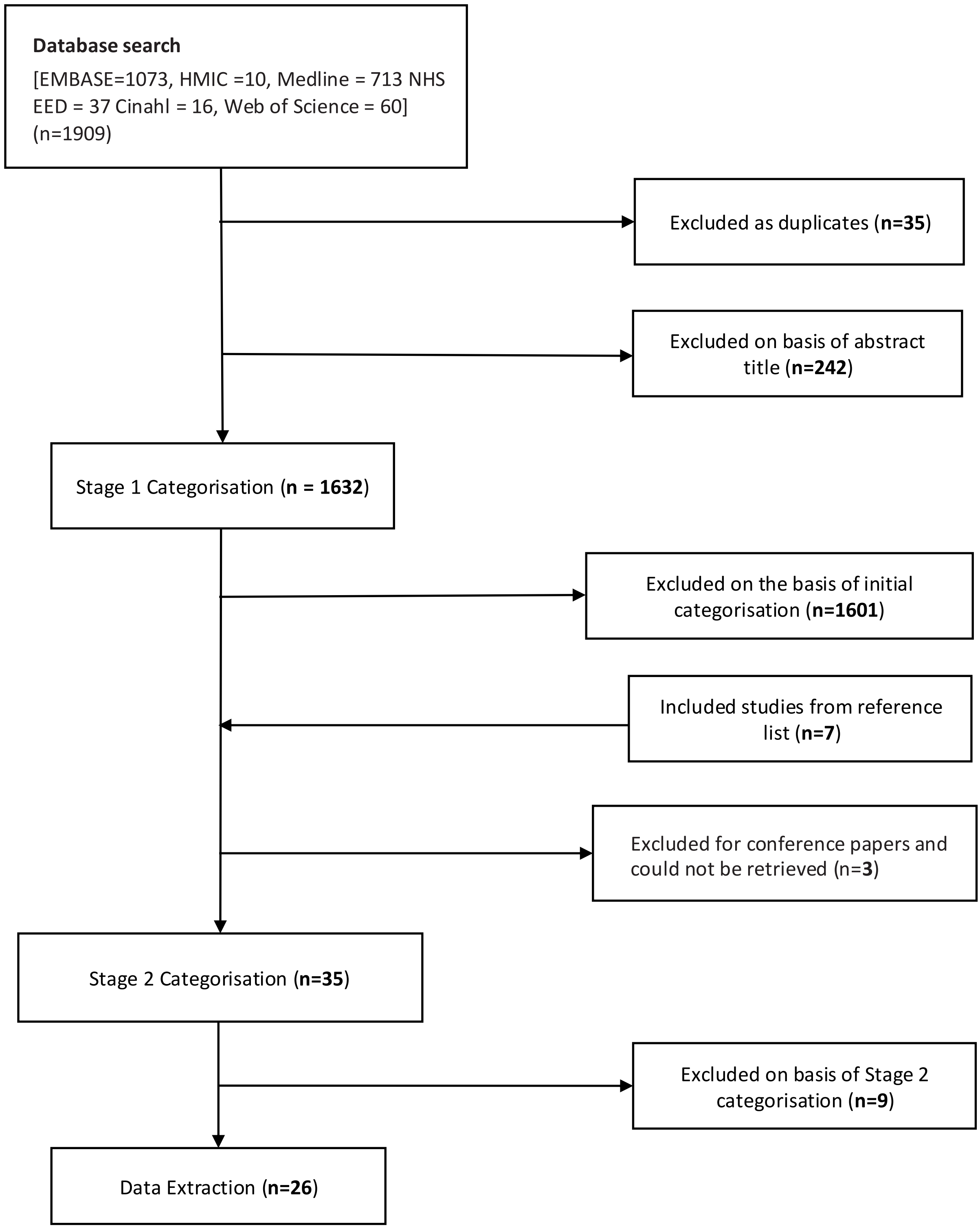


*S4: Figure 1 - Study selection*

*S5: Selected studies*

### Papers on PN


### Papers on Testing and Treatment

#### *S6: Studies on Testing and Treatment*

### *Patient Characteristics*

Fifteen papers reported various aspects of testing, screening and/or treatment of which fourteen studies were on MSM. The MSM studies focused on STIs (n=3)[12 15 18] HIV [5 11 13 14 16 17] (n=6) or both STIs and HIV (n=2).[6-10] The majority of the studies (9) used secondary data from published sources and national references, one study[26] used both primary and secondary data while the remaining studies used data collected alongside a trial[17] or from an STI Services.[13-15 18]

Evidence on patient characteristics was provided by a few papers. Three papers provided information on patients' age, of which two papers reported age groups as 13-64 years[5] and 15-64 years.[10] The third paper used a 30-year-old hypothetical patient who was followed for 80 years or until death. [9]

Six of the papers on MSM provided information on the setting of which four papers used primary data. Of these four papers, the first assessed data on patients attending an STI Clinic in Amsterdam.[18] The second study was a CUA alongside an RCT (eSTAMP Trial) in which patients were recruited through banner advertisements on sites typically used by MSM including internet social network, music and dating sites.[17]

The third study[14] assessed an MSM Testing Initiative (MTI) which implemented five HIV testing strategies. In liaison with pre-defined implementing partners, participants were sought by providing testing services at diverse settings such as (i) Public or private locations attended by MSM for purposes other than medical, mental health, or social, services. (ii) Couples voluntary counselling and testing (CVCT). (iii) Social network strategy (SNS) in which individuals encouraged peers to participate in HIV testing. (iv) Large-scale events including Gay Pride festivals, pageants and balls. Data were collected prospectively from implementing partners in 15 cities across the US.

The fourth study[13] collected data retrospectively from six Community Based Voluntary Counselling and Testing (CBVCT) services across Europe. There was no information on the recruitment method and the study provided the characteristics of testing and counselling centres rather than patients. [13]

Vriend et al.,[7] assessed screening for chlamydia in MSM with HIV and used MSM 'in care' at HIV treatment centres. The last paper obtained data from an established US database, which included data on men who have employment-based health insurance.[10] The authors assumed that the MSM population represents 3% of the total men. The remaining MSM studies did not provide any additional information on patients' characteristics. None of the papers reported on the type of partners.

### *Economic Evaluations and outcomes*

The 14 studies on MSM included nine CUAs, one CUA/CEA[18], one CEA[13] and three cost analysis,[9 10 15]. The CUAs used transmission dynamic Models,[5 7 12] Markov Models[6 8] or economic/ mathematical model[4 11 16] and reported outcomes in terms of cost per QALYs. The papers were not explicit about the reason for using a model, but it is assumed that the purpose was to see the impact on infection beyond the test, which is appropriate.

The majority of the model-based analyses used inputs including QALY weights from studies that were conducted on heterosexuals due to limited data on MSM. [5 6 8] However, a paper by Vriend et al.,[7] which assessed the cost-effectiveness of anorectal chlamydia screening among MSM in care at HIV treatment centres in the Netherlands, incorporated some QALYs from an MSM study[5] included in the current review (which had actually derived QALY weights from heterosexuals).

Zwart et al assessed the cost-effectiveness of testing strategies for Gonorrhoea amongst MSM attending an STI clinic.[18] Using inputs from a transmission model[18] the authors developed an economic (decision tree) model to estimate the long-term impact of the strategies on Gonorrhoea prevalence over 10 years. The authors reported outcomes as cost per case prevented and as cost per QALYs. The authors used HSUVs provided by a study by the Institute of Medicine in the United States.[29]

#### *Perspectives and costs*

There were only three MSM studies[5 6 14] that adopted the Societal perspective but two[5 6] of them reported only direct medical costs. The third paper[14] which also adopted a payer perspective assessed an MSM Testing Initiative (MTI) collected data for overhead costs such as equipment, set-up and implementation and staff costs. Eleven studies took the perspective of the healthcare system or the provider. While nine studies explicitly stated this, we made conclusions on four studies[10 11 13 17] based on the costs reported.

Shrestha et al.[17] assessed HIV testing in MSM alongside a trial in which participants were recruited through social media and dating sites that serve MSM and collected resource used for advertising, internet site design, monitoring for recruitment and incentives to participants.

Many of the studies collected data on consultations and office/GP visits. The studies reported total or average costs estimates for a range of activities including consultations, screening and testing (*Table 3*). Three studies which focused on testing[26] or treatment [9 18] also reported data on inpatient care/hospitalizations and outpatient care.

#### *S7: Characteristics of Selected Studies*

| # | First Author (Year) | Setting | Primary objective /Disease focus | Population/ Type | Method | Data Source | Intervention/Comparator |
|---|---|---|---|---|---|---|---|
| **Partner Notification** | | | | | | | |
| 1 | Katz (1988) | USA | Assess the efficiency and cost-effectiveness of using field follow-up for contacts of both untreated patients with chlamydia and at-risk individuals who were named contacts of men with NGU. **Chlamydia & NGU** | Individuals attending an STD clinic. Study 1: 142 patients (76 women & 66 men) Study 2: 678 men **Heterosexual** | CEA; Cost Analysis | Primary data from STD clinic | Study 1:Two different types of self-referral Study 2: Three methods of contacting female partners of men with NGU |
| 2 | Howell (1997) | USA | Evaluate the cost-effectiveness of 2 alternative PN strategies compared to no PN **Chlamydia** | A hypothetical cohort of 1000 men and 1000 women **Heterosexual** | CEA with Decision Model | Secondary data | 2 PN strategies: 1: locating and providing treatment for the female sex partners of male IPs; 2: locating and providing treatment for the male sex partners of female IPs vs No PN |
| 3 | Rahman (1998) | Japan | Assess the cost-effectiveness of a PN Program (PNP) **HIV** | 277 HIV carriers from the general population in 1995 **Heterosexual** | CEA with Analytic Model | Secondary data | Implementing PNP vs No PNP |
| 4 | Varghese (1999) | USA | Evaluate the cost-effectiveness of PN and counselling and testing offered in HIV and STD clinics in preventing future HIV infections **HIV** | A cohort of 10,000 individuals **Heterosexual** | CEA with Decision tree Model | Secondary data | PN vs Counselling and testing |
| 5 | Reynolds (2001) | USA | Assess the cost-effectiveness of selective screening compared with the strategy of PN in the detection of early syphilis. **Syphilis** | 2849 men and women with early syphilis **Heterosexual** | CEA | Primary data from a Sexual Health facility | Selective screening vs PN |
| 6 | Cohen (2004) | USA | Estimate the relative cost-effectiveness for 26 HIV prevention interventions including individual, biomedical, structural, and community & Social Network interventions. **HIV** | **Not stated MSM, Heterosexuals** | CEA using a spreadsheet tool developed with the Bernoulli process model | Secondary data from published sources | 26 prevention strategies including PN, testing & treatment and Mass Media Campaigns. |

| # | First Author (Year) | Setting | Primary objective /Disease focus | Population/ Type | Method | Data Source | Intervention/Comparator |
|---|---|---|---|---|---|---|---|
| 7 | Gift (2006) | USA | Evaluate the cost-effectiveness of the existing chlamydia screening and PN programs for men compared with 3 hypothetical alternatives. **Chlamydia** | A hypothetical cohort of 1000 male inmates **Not specified** | CEA; Decision tree model | Primary and secondary data | Age-based screening and testing based on symptoms of bacterial STDs vs existing program |
| 8 | Gunn (2006) | USA | Evaluate the usefulness of a syphilis model PN service for high-risk persons with chronic HBV infection. **HBV** | 129 patients aged 15-45 years (47 MSM; 26 IDUs & 12 MSM/IDUs **MSM** | Cost Analysis | Primary and Secondary data | PN services |
| 9 | Turner (2011) | UK | Compare cost, cost-effectiveness, and sex equity of different intervention strategies within the English NCSP. **Chlamydia** | Individuals aged 15-24 years **Heterosexual** | CEA; Economic & Mathematical Model | Secondary data | PN vs Screening |
| 10 | Roberts (2012) | UK | Compare costs and outcomes of two models of PN with routine patient referral PN, for sex partners of people with chlamydia, gonorrhoea and non-gonococcal urethritis to obtain cost data for APT strategies to use these data in a preliminary economic evaluation **Chlamydia** | Men and women **Heterosexual** | CCA | Primary & secondary data | Accelerated Partner Therapy (APT Hotline; APT Pharmacy) vs Routine PN |
| 11 | Nichols (2015) | The Netherlands | Determine the preventative impact on new HIV-1 infections and cost-effectiveness of PN **HIV** | Individuals aged 15 years and over **MSM** | CUA; Mathematical Model | Secondary/Local data | 2 PN scenarios assuming one will diagnose 5% and the other 20% |
| **Testing/Screening and Treatment** | | | | | | | |
| 1 | Postma (2001) | The Netherlands | Assess the cost-effectiveness of pharmacotherapy for male partners in screening women for asymptomatic infection with Chlamydia trachomatis **Chlamydia** | Heterosexually active young women (selective) visiting the general practices **Heterosexual** | CEA with Decision analysis Model | Primary (Amsterdam Pilot Study) and secondary data | Pharmacotherapy for: male partners with reinfection vs male partners without reinfection |
| 2 | Juusola (2011) | USA | Estimate the effectiveness and cost-effectiveness of strategies to identify acutely infected individuals and provide them ART **HIV** | Individuals aged 13-64 years. **MSM** | CUA; Transmission dynamic model | Secondary data/ Published literature & National references | 3 approaches: viral load testing for individuals with flu-like illness, expanded screening with antibody testing or viral load testing. |

| # | First Author (Year) | Setting | Primary objective /Disease focus | Population/ Type | Method | Data Source | Intervention/Comparator |
|---|---|---|---|---|---|---|---|
| 3 | Linas (2012) | USA | Estimate the effectiveness and cost-effectiveness of screening for acute hepatitis C virus (HCV) MSM with HIV **HIV & HCV** | A simulated cohort of HIV-infected MSM **MSM** | CUA; Monte Carlo Simulation Model | Secondary data | Screening and treatment strategies for HCV |
| 4 | Vriend (2013) | The Netherlands | Estimate the cost-effectiveness of anorectal chlamydia screening among MSM in care at HIV treatment centres. **HIV and Anorectal Chlamydia** | **MSM** | CUA; Transmission Model | Secondary data | Biannual screening vs Annual screening |
| 5 | Tuite (2014) | Canada | Evaluate if enhanced routine (more frequent and/or higher coverage) syphilis screening in HIV-positive MSM is more cost-effective relative to the current standard of care **HIV and Syphilis** | **MSM** | CUA; Individual-level Microsimulation Model (Markov Model) | Local reports/database | Higher coverage vs Usual care at specified time points |
| 6 | Nakagawa (2015) | UK | Calculate the expected lifetime HIV-related healthcare cost for HIV infection and explore the extent to which the lifetime costs would be reduced if patented drugs were replaced by generic antiretroviral drugs. **HIV** | **MSM** | Cost Analysis; Stochastic Computer Simulation Model | Secondary data/Published sources | Generic ART vs Patented ART |
| 7 | Owusu-Edusei (2015) | USA | Estimate the annual total direct medical cost of providing the recommended STI and HIV testing and counselling services for all MSM **STI/HIV** | Individuals aged 15-64 years **MSM** | Cost Analysis | Secondary data | STI and HIV testing and counselling services |
| 8 | Hutchinson (2016) | USA | Assess the cost-effectiveness of various testing strategies and timelines for MSM and IDU. **HIV** | Cohorts of MSM and IDUs **MSM** | CUA; Mathematical model | Secondary data | 2 HIV Testing Strategies at 3- and 6-month intervals vs annual testing |
| 9 | Xiridou (2016) | The Netherlands | Explore if dual therapy will delay the spread of antibiotics resistance for gonorrhoea compared to monotherapy. **Gonorrhoea** | **MSM** | CUA; Transmission dynamic model | Secondary data | Dual Therapy (Ceftriaxone plus Azithromycin) vs Monotherapy (Ceftriaxone) |

| # | First Author (Year) | Setting | Primary objective /Disease focus | Population/ Type | Method | Data Source | Intervention/Comparator |
|---|---|---|---|---|---|---|---|
| 10 | Perelman (2017) | Six Communities in 5 European Countries (Greece, Denmark, Portugal, Slovenia & France) | Conduct an economic evaluation of the activities of six European Community Based Voluntary Counselling and Testing (CBVCT) services **HIV** | **MSM** | CEA; Cost Analysis | Primary data retrospectively from CBVCT Services | Six CBVCT Services |
| 11 | Zulliger (2017) | USA | Evaluate the costs and cost-utility of 3 HIV testing strategies **HIV** | 27,475 Particpants **MSM** | CUA; Cost Analysis | Primary data from 16 implementing partners in 15 cities | Testing strategies |
| 12 | Bartelsman (2018) | The Netherlands | Estimate the impact of testing on prevalence and costs of testing and treatment for anogenital gonorrhoea. **Gonorrhoea** | **MSM** | Cost Analysis; Transmission dynamic model | Secondary data | Point of Care (POC) for Symptomatic MSM only or all MSM vs No POC for MSM |
| 13 | Reitsema (2019) | The Netherlands | Assess the cost-effectiveness of increased consistent HIV testing **HIV** | **MSM** | CUA; Stochastic Economic Model | Secondary data | HIV testing rates |
| 14 | Shrestha (2020) | USA | Estimate the costs and cost-effectiveness of delivering a self-testing programme **HIV** | **MSM** | CUA; Cost Analysis | Primary data from an RCT | HIV self-testing vs No self-testing only access to Trial (eSTAMP) Website |
| 15 | Zwart (2020) | The Netherlands | Assess the cost-effectiveness of three testing strategies for microscopic Gram-stained smear (GSS) evaluation for the detection of anogenital gonorrhoea **Gonorrhoea** | **MSM** | Model-based CUA/CEA | Primary data from STI clinic | No GSS and GSS in symptomatic and asymptomatic MSM vs GSS in symptomatic MSM only |

#### *S8: Selected results*

| | Author (Year) | Perspective | Cost type | Resource use and costs | Year and currency | Key costs | Primary outcome |
|---|---|---|---|---|---|---|---|
| **Partner Notification** | | | | | | | |
| 1 | Katz (1988) | N/S | Direct & Indirect Medical costs | Staff time<br>Salaries<br>Phone calls<br>Postage<br>Travel | N/S<br>USD | Untreated infected patients<br>- Field follow-up ($13.52 to $20.06)<br>- Reminder system ($21.27 to $67.05)<br>Contacts of NGU<br>-Nursing referral: $42.46<br>-Interview only: $60.48<br>- Field trip: $37.50 | Number of partners elicited<br>Number of partners treated |
| 2 | Howell (1997) | Healthcare System | Direct program costs for PN<br>Direct Medical costs for Chlamydia treatment | Inpatient care<br>Outpatient care | 1994<br>USD | Sequelae costs - $567,000<br>PN interview with staff -$23.72<br>Cost of locating one partner - $35.75<br>Case of PID - $4890.68<br>Medication (Azithromycin) - $9.50 | Cases of PID prevented<br>Cost per Index patient (IP) |
| 3 | Rahman (1998) | N/S but probably Healthcare provider | Direct costs | Counselling of index cases<br>Locating partners;<br>Counselling and testing of partners:<br>Initial check-up, follow-up, Treatment of the newly found HIV carriers and others | 1997<br>USD | Follow-up and antiviral treatment of newly found HIV carriers $ 1.8 million,<br>Treatment of AIDS till death $ .69<br>Medical care during the gained life years $ 0.48 million<br>Ancillary care $0.49million<br>Locating, counselling and testing of the partners (including counselling of index cases) $ 0.18 million | Cost per life year saved |
| 4 | Varghes e (1999) | Societal & Provider | For Societal - Costs incurred by providers and clients<br>For Provider – Direct Medical costs | N/S | 1997<br>USD | Cost estimates for counselling and testing and partner notification programs and lifetime treatment cost of HIV | Cost per case prevented |
| 5 | Reynolds (2001) | Health Department | Direct Medical & Non-Medical costs | Staff wages<br>Staff time - Duration of case management, lab tests & paperwork<br>Travel | 1996<br>USD | Selective screening (Testing; Contacting infected patients to return for treatment)<br>Selective screening ($395)<br>PN Management (Surveillance; case management; serologic testing of sexual contacts and prophylactic treatment activities) PN ($405) | Cost per case of early syphilis detected;<br>Selective screening was more cost-effective with: ICER of $239 Primary cases; ICER of $553 of Secondary cases |

| | Author (Year) | Perspective | Cost type | Resource use and costs | Year and currency | Key costs | Primary outcome |
|---|---|---|---|---|---|---|---|
| 6 | Cohen (2004) | Public Health System | Direct Medical & indirect | Salaries<br>Staff time<br>supplies<br>overhead | USD<br>N/S | Cost per Person hours<br>Cost to intervene with an individual person at risk<br>Cost for the entire community divided | Number of HIV infections prevented<br>person reached by intervention<br>cost per person reached by the intervention<br>cost per new HIV infection prevented |
| 7 | Gift (2006) | 2 perspectives: Correctional institution and Healthcare system. | Direct Medical & Indirect | Number tested<br>Number treated<br>Correctional perspective: program costs - overhead, labour, testing, treatment, and sequelae and overhead costs.<br>Healthcare system perspective: testing and treatment, sex PN program and costs for treating female partners and PN Services cost | 2001<br>USD | Healthcare System cost – 17,670 to 39,820<br>Net PN cost – 90 to 800 (symptom based to universal screening) | Cost per case treated – 503 (Age based <30 yrs.) to 1591 (symptom based)<br>Cases of PID averted in female sex partners as primary cost-effectiveness outcomes<br>Universal screening most effective and most costly with ICER of 4592 per cases averted |
| 8 | Gunn (2006) | N/s | N/S | Number of phone calls (10 minutes) & field visits (120 minutes)<br>Staff hours | N/S | Serology costs - $510<br>Vaccine costs - $903<br>Total PN services cost - $20,613, or $1472 per vaccine | PN services cost per vaccine |
| 9 | Turner (2011) | Healthcare Provider | Direct Medical costs | PN – Duration of PN Consultations or clinician delivered treatment; provider referral by health professionals,<br>and follow-up telephone calls .<br>Screening - number screened, number infected, | 2008-2009<br>GBP | cost of a screen - £43.65 (£32.01 to £54.32)<br>cost per individual tested,<br>cost per positive diagnosis,<br>total cost of screening | Cost of PN per positive index case (£2 to £13)<br>Screening costs per infected individual treated £506 (£381–£621) |

| | Author (Year) | Perspective | Cost type | Resource use and costs | Year and currency | Key costs | Primary outcome |
|---|---|---|---|---|---|---|---|
| 10 | Roberts Num(2012) | NHS | Direct Medical | Intervention (mobile phone & credit)<br>Duration of consultation Uptake of treatment pack<br>Intervention<br>Admin work | 2008<br>GBP | Cost of a sexual health check-up for the routine arm - £45.89.<br>Routine PN (£46) APT (£54) APT Pharmacy (£53) | Average Cost per partner treated |
| 11 | Nichols (2015) | Third party payer | Direct medical costs | Hospitalization opportunistic infections, HIV testing, and ART | 2015<br>Euro | Primary HIV test† €20Confirmatory testing €45 STI tests (chlamydia, gonorrhea, syphilis, hepatitis B and HIV combined)<br>€124<br>Outpatient visit at clinic/primary care €31<br>Outpatient visit after diagnosis €124<br>Outpatient visit for partner notification €124 , Local data<br>Outpatient visit at HIV specialist €143<br>Cost of treating opportunistic infections, Recent infection €4.47-€21.62 | QALY<br>PN Cost effectiveness:<br>5 years - €41,736 per QALY<br>20 years - €5,887 per QALY |
| **Testing/Screening and Treatment** | | | | | | | |
| 1 | Postma (2001) | Societal | Health care costs and productivity losses | Inpatient care<br>Outpatient care | 1996<br>Euro | Investment (€1,227,100 to €1,185,700)<br>Savings (€638,000 to €1,033,700)<br>Net costs (€152,000 to €547,700)<br>Outcomes averted (€701 to €1,153)<br>Cost-effectiveness ratio (€132 to €781) | Averted cases of PID and infertility (major outcomes).<br>Reinfection in the absence of partner pharmacotherapy<br>Net costs per major outcome averted |
| 2 | Juusola (2011) | Societal (but only health-related costs were reported)) | Medical costs | HIV healthcare costs, cost of ART, cost of HIV testing protocols diagnosis and Counselling | 2009<br>USD | Annual HIV-related healthcare costs<br>Cost of HIV testing-VL test<br>Cost of HIV testing-antibody test<br>Cost of counselling<br>Cost of HIV diagnosis | Cost per HIV case<br>Cost per QALY<br><br>ICERs: Current screening rates $115,325 per QALY<br>Expanded screening coverage $105,398 per QALY |
| 3 | Linas (2012) | Societal (reported only medical costs) | Medical costs | N/S | 2011<br>USD | Cost of screening testing<br>Cost of confirmatory testing<br>Cost of HCV therapy/month | QALY<br>**ICER values**<br>NEAT-recommended screening, compared with symptom based |

| | Author (Year) | Perspective | Cost type | Resource use and costs | Year and currency | Key costs | Primary outcome |
|---|---|---|---|---|---|---|---|
| | | | | | | | screening, was $43,700/QALY gained<br>3-month LFTs compared with NEAT was $129 700/QALY gained |
| 4 | Vriend (2013) | Healthcare Provider | Medical cost | Uptake of chlamydia screening, Treatment of<br>three infections( HIV, symptomatic chlamydia, and asymptomatic chlamydia),<br>Referral to an STI clinic for further testing, counseling, and PN. | 2009<br>Euro | **Costs of new screening program:**<br>Testing for chlamydia among MSM in care at an HIV treatment center €59<br>Treating chlamydia (antibiotics) and referral to STI clinic when found in MSM in care at an HIV treatment center €200<br>**Costs outside new screening program:**<br>STI testing (mean over GP, STI clinic and dermatology) €203<br>Treating chlamydia (mean over GP, STI clinic and dermatology) €7<br>Treating epididymitis (complication of chlamydia) €325 | Averted HIV and chlamydia infections<br>QALY |
| 5 | Tuite (2014) | Healthcare Provider | Direct medical costs | Testing, treatment, follow-up , lifetime costs for untreated Individuals and adverse event management (GP mainly). | 2011<br>CAD | Diagnostic tests<br>Treatment<br>Adverse events | QALY<br>Life expectancy<br>Higher coverage,<br>6 months was associated with more QALYs and lower cost compared to other strategies except for higher coverage 3 months which was more costly yet more effective with an ICER of CD $77,516 |
| 6 | Nakaga wa (2015) | Healthcare Provider | Direct medical costs | Healthcare Centre visits for 3 month period, inpatient and outpatient care | 2013<br>GBP | Use of healthcare centre services (weighted average for inpatient, outpatient and day ward)<br>Cost of testing<br>Cost of treatment | Life expectancy<br>mean lifetime cost of HIV treatment<br>- non-discounted £360,800<br>- Discounted £185,200 |
| 7 | Owusu-Edusei (2015) | N/S but probably Healthcare Provider) | Direct medical costs | STI and HIV tests, counseling and office visit | 2014<br>USD | **Average cost/service**<br>Chlamydia (rectal, urethral) $45 ; HBV $23;<br>High Intensity Behavior Counseling $29; | N/A<br>Annual cost of providing the complete set of STI and HIV |

| | Author (Year) | Perspective | Cost type | Resource use and costs | Year and currency | Key costs | Primary outcome |
|---|---|---|---|---|---|---|---|
| | | | | | | HIV $18; HSV-2 $27; Gonorrhea (pharyngeal, rectal, urethral) $45<br>Office visit $100 ; Syphilis $8 | testing and counseling for all MSM was $1.1 billion |
| 8 | Hutchinson (2016) | N/S but probably Healthcare Provider | laboratory costs<br>costs for client recruitment & outreach<br>Cost of lifetime treatment | Testing | 2012<br>USD | **HIV cost per test /per-person**<br>Rapid HIV-; HIV + $22.62; $98.32<br>Fourth-generation IA HIV-; HIV + $10.84; $73.51<br>Client recruitment + outreach $16.07<br>Lifetime HIV treatment costs $417,000<br>Annual treatment costs $16 | QALY<br>HIV infection averted<br>ICER $48,000/QALY |
| 9 | Xiridou (2016) | Healthcare Provider | Medical costs | Duration of Consultations at STI clinics and GPs<br>- Tests<br>- doses of ceftriaxone, azithromycin, and alternative antibiotics (ciprofloxacin or amoxicillin) | 2012<br>Euro | GP consultation €29.73<br>STI clinic consultation €29<br>NAAT test €35.18<br>Culture test (for 4–6 media) €36.24<br>Treatment with ceftriaxone (500 mg 1 injection) €4.19<br>Treatment with azithromycin (2 tablets of 500 mg) €0.41<br>Treatment with alternative antibiotic €0.24<br>Pharmacy handling fee per receipt item €12<br>Pharmacy handling fee per item delivered to STI clinics €0.12 | QALY<br>ICER of dual therapy compared to monotherapy |
| 10 | Perelman (2017) | N/S but probably Healthcare Provider | Medical cost<br>Financial expenditure to deliver the services during the 12-month period | Facility, professional and voluntary staff, testing material, equipment, transportation, first visit and related exams, communication | 2014<br>Euro | Facility €1,080-54,123<br>Professional staff €12,883-146,012<br>Voluntary staff €0-100,444<br>Testing material €10649-46599<br>Equipment €0-2866<br>Transportation €0-2352<br>First visit and related exams €0-412<br>Communication €2016-13473<br>Other administrative €779-9408<br>Total cost: €54390-245803<br>Only lower and upper costs were extracted as the study reported costs for cities | cost per HIV test<br>cost per reactive HIV test<br>Total costs of CBVCTs: 54,390€ to 245,803€ per year<br>Cost per HIV test €41-113<br>-cost per reactive HIV test €1966-9065<br>Cost per reactive HIV linked to care €2297-20215 |
| 11 | Zulliger (2017) | Payer & Societal | Direct Medical & Indirect costs | Total implementation cost; Staff costs | 2013-14<br>USD | Total implementation cost $3,085,500<br>Staff costs $1,738,592 | Cost per QALYs gained<br>Cost per a new HIV diagnosis |

| | Author (Year) | Perspective | Cost type | Resource use and costs | Year and currency | Key costs | Primary outcome |
|---|---|---|---|---|---|---|---|
| | | | | Overhead | | | |
| 12 | Bartelsman (2018) | Healthcare Provider | Direct Medical costs | Direct staff time (salary plus benefits),<br>supplies,<br>Overheads<br>Treatment | 2008-2011 Euro | **Cost per consultation**<br>Initial visit €12.11 (100% staff)<br>Culture test €13.15 (37% staff, 15% supplies, 48% culture<br>Gram test €3.71 (81% staff, 19% supplies)<br>Treatment €11.44 (87% staff, 13% supplies)<br>Gram negative result €0.81 (100% staff)<br>Patient follow up for negative result €2.42/attempt (100%staff) | **Number of treated infection**<br><br>**Cost per consultation** €26.24 to €30.47<br>**Cost per correctly managed consultation** €26.38 to €31.12<br>**Cost per treated infection** €320.44 **to** €495.22 |
| 13 | Reitsema (2019) | Healthcare Payer | Medical costs | Consultations, laboratory tests, HIV-medication, dispensing fees for pharmacies, treatment of opportunistic infections, and treatment of gonorrhea and epididymitis | 2016 Euro | Costs HIV test<br>Costs of monitoring per HIV-positive individual in care<br>cART costs per person<br>Costs of treatment of opportunistic infections<br>Annual hospitalization per person €7103.5 | QALY<br>ICERs<br>Small increase in testing MSM €27931/QALY<br>Moderate MSM €36694/QALY |
| 14 | Shrestha (2020) | Healthcare Provider (Not clear) | | Internet site design<br>Monitoring for recruitment,<br>programme administration, and overhead, HIV test kits, mailing test kits, supplies, incentives to participants. | 2016 USD | **Advertising and recruitment cost/person interviewed**<br>Dating site $134.46 -m$221.88<br>Pandora internet radio site $78.31<br>Gay Ad Network $122; Facebook $39.82<br>MUSED magazine $2.36<br>**Other costs**<br>Cost per self-test completed $61<br>Cost per person tested $145<br>**Total costs of program implementation are also reported in the study** | QALY<br>HIV diagnosed<br><br>Cost per new HIV diagnosis (C/N) $9365<br>Cost per HIV transmission averted (C/A) $134,583<br>Cost per QALY saved (C-AT)/AQ ($74,476) |
| 15 | Zwart (2020) | Healthcare Payer | Direct Medical costs | Medication; GP visits; outpatient visits; inpatient visits | 2016 Euro | Testing costs;Total healthcare costs<br>Treatment costs for gonorrhoea<br>Complication-related healthcare costs | Number of epididymitis cases<br>QALY; ICER (€/prevented epididymitis case);ICER (€/QALY gained |
